# Divergence Between Net Fluid and Weight-Based Evaluation in Calculating Cumulative Fluid Balance

**DOI:** 10.1101/2025.10.22.25338544

**Authors:** Finley J. Shinnick, Denise C. Hasson, Ulka Kothari, Ami Shah, James D. Odum, Chloe G. Braun, Celeste G. Dixon, Julie C. Fitzgerald, Susan D. Martin, Nina Terry, Adam C. Dziorny, the Evidence-Driven Evaluation and Management of fluid Accumulation (EDEMA) Collaborative

## Abstract

**Objective:** Although efforts have been made to standardize fluid balance calculations in the intensive care unit (ICU), there is a limited understanding of how different calculation methods relate to one another across an ICU admission. We quantified the agreement between the cumulative fluid balance calculated from fluid intake and output (CFBf) and from serial weights (CFBw) in critically ill children during the first week of ICU admission.

**Design:** Retrospective, multicenter, federated observational study.

**Setting:** Four pediatric medical-surgical ICUs (PICU) and two pediatric cardiac ICUs (PCICU) from four tertiary care centers.

**Patients:** Analysis included 8,895 pediatric patients (<19 years old) representing 12,388 ICU encounters from 2023-2024.

**Interventions:** None.

**Measurements and Main Results:** A patient’s anchor weight was the weight closest to ICU admission. CFBf and CFBw were calculated at the time of new weight measurements. We assessed agreement between CFBf and CFBw using Bland-Altman analyses, stratified by ICU day and patient subgroups (neonates, early anchor weights [weight on ICU day 0], and encounters with unmeasured urine occurrences). Across all units and subgroups, CFBf exceeded CFBw (mean difference: all patients = 4.7 %CFB, early anchor weight = 4.7 %CFB, neonates = 5.9 %CFB). The mean difference increased significantly over time (days 0–3: 2.7% vs. days 4–7: 8.1%, p<0.05), with greater divergence in neonates and those with early anchor weights.

**Conclusions:** CFBf consistently exceeded CFBw across all subgroups, with a greater divergence on ICU days 4-7. Clinicians should understand these differences, prioritizing early and frequent patient weights throughout ICU admission. Future studies should assess each method’s association with patient outcomes to identify the most clinically informative CFB method.

**Research in Context:**

- Cumulative fluid balance (CFB) is calculated by fluid (intake minus output) and weight measurements in pediatric ICU patients, but few studies have directly compared these methods across multiple institutions, unit types, and throughout PICU admission.
- CFB calculated using fluid measures consistently exceeded weight-based calculations, with divergence increasing later in admission and for patients with anchor weights recorded closer to PICU admissions and neonates.
- Divergence between CFB methods may reduce CFBf reliability later in the ICU stay, so accurate and frequent PICU weight recordings are essential for reliable fluid balance assessment.

**At the Bedside:**

- This multicenter study demonstrates a consistent positive divergence between CFB calculated by net fluid compared to weight-based calculations across diverse PICU and PCICU populations.
- Differences between methods diverged later in admission (e.g., after day 3), particularly among neonates and patients with earlier recorded anchor weights, while CFB calculated before the first unmeasured urine differed little from the overall cohort.
- Clinicians should understand these differences and the factors that influence them, prioritizing early and frequent patient weights throughout ICU admission to recognize when net fluid-based CFB diverges from a weight-based CFB.

## INTRODUCTION

Fluid overload, or the pathologic increase in cumulative fluid balance (CFB), occurs in more than one-third of children admitted to the pediatric intensive care unit (PICU) (1) and is associated with multiple morbidities (2). Due to the dose-dependent association between fluid overload and poor outcomes in PICU populations, precise tracking of CFB across the course of admission is essential for guiding clinical decision-making (3–7). Although efforts have been made to standardize fluid balance definitions and calculations in the PICU (8), there is a limited understanding of how different calculation methods relate to one another across admission (9,10).

Two commonly accepted methods are used to assess CFB throughout ICU admission, each with its own advantages and disadvantages (11). The first method compares net fluid intake and output relative to an anchor weight (commonly the ICU admission weight) (CFBf) (12). While this method aims to capture fluid that remains in a patient, there is uncertainty in quantifying unmeasurable fluid losses (e.g., insensible losses), and it is prone to error when certain outputs cannot be measured (e.g., flushed urine, emesis, unweighed stool) (13). The second method calculates CFB by serial weight changes (CFBw) relative to an anchor weight, which assumes that all weight change is solely attributable to fluid accumulation (14). Weighing critically ill children can be cumbersome, resource-intensive, and inaccurate due to lines, drains, and tubes, and it is often not done routinely. Both methods require an anchor weight, which serves as the reference point for changes in fluid volume or weight, but which is not standardized across units, centers, and is not always performed on ICU admission. Despite limitations, each method aims to capture a patient’s true CFB, yet their degree of agreement beyond the first 48 hours of admission across diverse patient populations remains unclear.

The objective of this multi-institutional study was to quantify the agreement between CFBf and CFBw, stratified by patient populations over the first 7 days of PICU admission. We report the variability in obtaining daily weights across units and the general features of CFB evaluation by each method, and specifically how they differ in high-risk populations (e.g., neonates) and with different methodologies (e.g., definition of anchor weight). We hypothesize that CFBf and CFBw will diverge as a patient gets later into their ICU stay, and divergence will be greater in neonates and those in whom anchor weight is recorded after the first day of ICU admission.

## METHODS

### Study Design

We performed a retrospective observational cohort study within our Evidence-Driven Evaluation and Management of fluid Accumulation (EDEMA) collaborative (4, 15). This study followed procedures in accordance with ethical standards as delineated by the Helsinki Declaration of 1975. Each unit’s institutional review board (IRB) deemed the project exempt from IRB review.

Characteristics of our collaborative member units have been described previously (4). We included data from six PICUs within four tertiary care centers: 132 beds from four mixed medical/surgical units (PICUs) and 53 beds from two pediatric cardiac ICUs (PCICUs), totaling 185 beds. All units use the same electronic health record (EHR), Epic (Epic Systems Corporation, Verona, WI). Our patient cohort included all pediatric patients <19 years old admitted to the PICU/PCICU between each unit’s respective date ranges, generally spanning January 1, 2023, to July 1, 2024. (**Table 1**). Similar to our collaborative’s prior federated studies, each unit maintained the privacy of its data, but identical data structures allowed for the sharing of data queries and analysis scripts (4).

**Table 1.** Patient demographics and clinical characteristics for each unit. The date range indicates the period of data extraction for each unit. Percentages for patient demographics indicate the proportion of patients in a given demographic at a given unit. Cumulative ICU days include the sum of all encounters’ days 0-7 in the ICU. Frequency of additional weights gives the count of additional weights and the ratio to each unit’s total ICU encounters (e.g., in Unit A, 2,351 total additional weights were measured beyond each encounter’s anchor weight. This was divided by 1,902 ICU encounters to yield 1.2 additional weights per ICU encounter). The additional weight for ventilated patients shows how many extra weights were taken for patients on a ventilator and what percentage of all additional weights that represents. Encounters with anchor weights after day 0 are those whose anchor weight was taken after ICU day 0, and proportions relative to total encounters at each unit. Encounters with duplicated weights had additional weights that varied by less than 0.001 kg from their anchor weight. ICU-intensive care unit, yr-year, wt-weight.

| Unit | Unit A | Unit B | Unit C | Unit D | Unit E | Unit F |
| --- | --- | --- | --- | --- | --- | --- |
| ICU Type | Cardiac ICU | Cardiac ICU | PICU | PICU | PICU | PICU |
| ICU Encounters, n | 1869 | 631 | 715 | 6266 | 1198 | 1509 |
| Female, n (%) | 823 (43%) | 285 (45%) | 307 (43%) | 2655 (42%) | 531 (44%) | 688 (45%) |
| Age, median (IQR) | 0.2 (0, 2.6) | 0.4 (0.01, 1.8) | 7.1 (1.8, 7.9) | 4.0 (0.9, 11) | 6.1 (1.8, 7.7) | 5.5 (1.5, 13) |
| Weight (kg), n (%) |  |  |  |  |  |  |
| < 10kg | 1092 (57%) | 327 (52%) | 146 (20%) | 1353 (21%) | 241 (20%) | 359 (24%) |
| 10-30 kg | 515 (27%) | 175 (28%) | 271 (38%) | 2964 (47%) | 473 (39%) | 605 (40%) |
| >= 30 kg | 262 (14%) | 129 (20%) | 298 (42%) | 1949 (31%) | 484 (40%) | 545 (36%) |
| Race, n (%) |  |  |  |  |  |  |
| White | 899 (47%) | 334 (53%) | 401 (56%) | 2671 (42%) | 712 (59%) | 893 (59%) |
| Black or African American | 278 (15%) | 59 (9%) | 248 (35%) | 1697 (27%) | 245 (20%) | 225 (15%) |
| Asian | 73 (4%) | 23 (4%) | 2 (0.28%) | 272 (4%) | 26 (2%) | 83 (5%) |
| Native Hawaiian or Pacific Islander | 1 (0.05%) | 1 (0.16%) | 0 (0%) | 0 (0%) | 0 (0%) | 5 (0.33%) |
| American Indian or Alaskan Native | 10 (0.53%) | 1 (0.16%) | 2 (0.28%) | 4 (0.06%) | 1 (0.08%) | 12 (0.79%) |
| Other/Multiple | 497 (26%) | 119 (19%) | 24 (3%) | 1480 (23%) | 162 (14%) | 104 (7%) |
| Unknown/Refused | 111 (6%) | 94 (15%) | 38 (5%) | 140 (2%) | 51 (4%) | 187 (12%) |
| Ethnicity, n (%) |  |  |  |  |  |  |
| Hispanic/Latino | 416 (22%) | 35 (6%) | 67 (9%) | 1092 (17%) | 112 (9%) | 221 (14%) |
| Non-Hispanic/Latino | 1327 (70%) | 412 (65%) | 604 (84%) | 4956 (78%) | 920 (77%) | 495 (32%) |
| Other/Unknown | 126 (7%) | 184 (29%) | 44 (6%) | 216 (3%) | 166 (14%) | 793 (52%) |
| Clinical Characteristics |  |  |  |  |  |  |
| Date Range | 01/01/2023 - 07/01/2024 | 01/01/2023 - 07/01/2024 | 12/01/2023- 05/01/2024 | 01/01/2023 - 07/01/2024 | 01/01/2023 - 07/01/2024 | 01/01/2023 - 07/01/2024 |
| Cumulative ICU Days | 9918 | 3414 | 2635 | 27370 | 4609 | 6311 |
| Encounters | 1869 | 631 | 715 | 6266 | 1198 | 1509 |
| Median Length of Stay [IQR], days | 4 [2, 6] | 5 [2, 7] | 2 [1, 3] | 3 [2, 5] | 2 [1, 4] | 2 [2, 4] |
| Frequency of Additional Wt., n (ratio to ICU encounters) | 2352 (1.3) | 1131 (1.8) | 1321 (1.8) | 3260 (0.5) | 231 (0.2) | 733 (0.5) |
| Additional Wt. for Ventilated Patients | 1544 (65%) | 267 (24%) | 565 (43%) | 1636 (50%) | 126 (55%) | 466 (64%) |
| Encounters with Anchor Wt after Day Zero, n (%) | 183 (10% %) | 35 (6%) | 37 (5%) | 359 (6%) | 22 (2%) | 66 (4%) |
| Encounters with Urine Occurrences | 324 (17%) | 496 (79%) | 261 (37%) | 2720 (43%) | 828 (69%) | 540 (36%) |
| Encounters with Duplicated Wt. | 93 (5.0%) | 53 (8.4%) | 169 (2.4%) | 202 (3.2%) | 73 (6.1%) | 107 (7.1%) |

### Measures and Outcomes

We collected baseline demographics, patient weights, total intake and output fluid volume by route and subtype, and the number of unmeasured urine occurrences. Consistent with prior studies, we defined Day 0 of ICU stay as admission time through 7 AM the following day (2). We defined Day 1 as the subsequent 24 hours (7 AM to 7 AM), and other days in the following 24 hours until Day 7 or the patient’s end of ICU admission. We defined anchor weight as the measurement temporally closest to ICU admission, whether obtained before or during ICU admission. Due to the federated nature of this study, we could not distinguish potential bias between pre- and post-admission weights. If a patient’s anchor weight was recorded before ICU admission, we considered that weight to be the one used for ICU clinical decision-making, and we used that weight for all calculations.

Acknowledging the variability in timing of weights obtained during ICU admission, we calculated CFBw at the time of each additional recorded weight as the change from the anchor weight, divided by the anchor weight. At each non-admission weight measurement timestamp, we calculated CFBf as cumulative fluid intake minus output divided by anchor weight (Equation 1). Granularity of fluid intake and output measurements was to the day, while weights were reported as seconds since ICU admission. To identify fluid values at specific weight timestamps, we proportionally distributed each subtype of fluid intake and output over the ICU day, and we calculated net fluid intake and output relative to anchor weight (Equation 2). We also calculated a truncated CFBf (Pre-Occurrence), to exclude any CFB value that followed a patient’s first unmeasured urine occurrence.

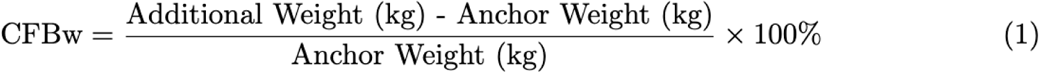

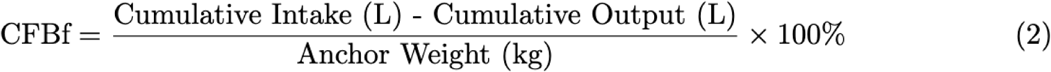

We conducted subgroup analyses for patients who had a weight measurement taken on their ICU day 0 (Early Anchor Weight) and patients younger than one month (Neonate). Early anchor weight allows a truer assessment of changes in CFB over the full ICU admission, while neonates, who typically have smaller body weights and may experience nutritional weight gain, are more sensitive to the impact of changes in fluid volume or weight on CFB calculations. We also segregated all CFB calculations into ICU days 0-3 and ICU days 4-7 to understand the relationship between CFBf and CFBw throughout ICU admission.

### Statistical Analysis

Statistical analysis was performed using R (16). We used Shapiro-Wilk tests to assess normality between the mean difference between CFBf and CFBw in both early versus late ICU timeframes. We subsequently applied Whitney-Mann tests or Student t-tests to assess differences in the mean difference of CFB methods between time in ICU admission. We used linear models to assess unit variation in obtaining and recording weights, based on patient age group and ventilator status (e.g., active order placed for invasive ventilation). We calculated raw CFBf and CFBw values across admission to inform the general direction of the mean difference between methods (**Figure 1**). We performed Bland-Altman analysis to assess the relationship between CFBf and CFBw (mean difference +/- standard deviation). Bland–Altman plots for each unit across ICU days 0 – 7 showed greater scatter later in admission, prompting further evaluation of the relationship between CFBf and CFBw on ICU days 0 – 3 versus 4 – 7 (**Figure 2**). We could not compute regression analyses across units due to the federated nature of this study. We defined a significant difference as p<0.05.

**Figure 1:**
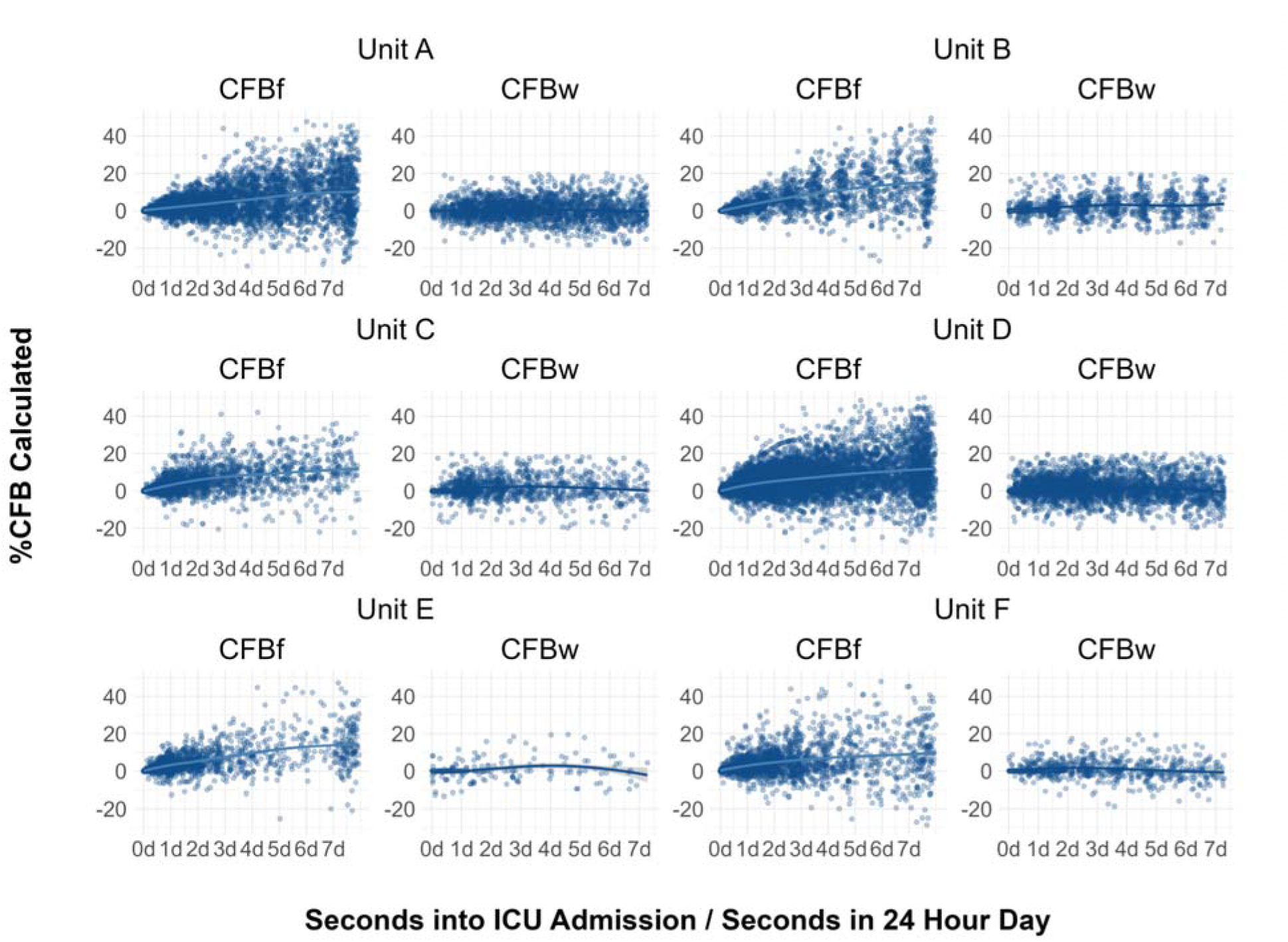
Graph showing CFB values calculated by weight and fluid method at each unit. Values plotted in seconds since ICU admission, divided by seconds in 24 hours to approximate days in ICU admission. A local polynomial regression is shown for each plot to visualize trends. Values for CFBf were calculated at times when a weight measurement was recorded in addition to the time patients were discharged from the ICU. CFBw values are only shown at non-anchor weights. Values <-30% or >50% are not graphed for visual clarity, but were included in the analysis.

**Figure 2:**
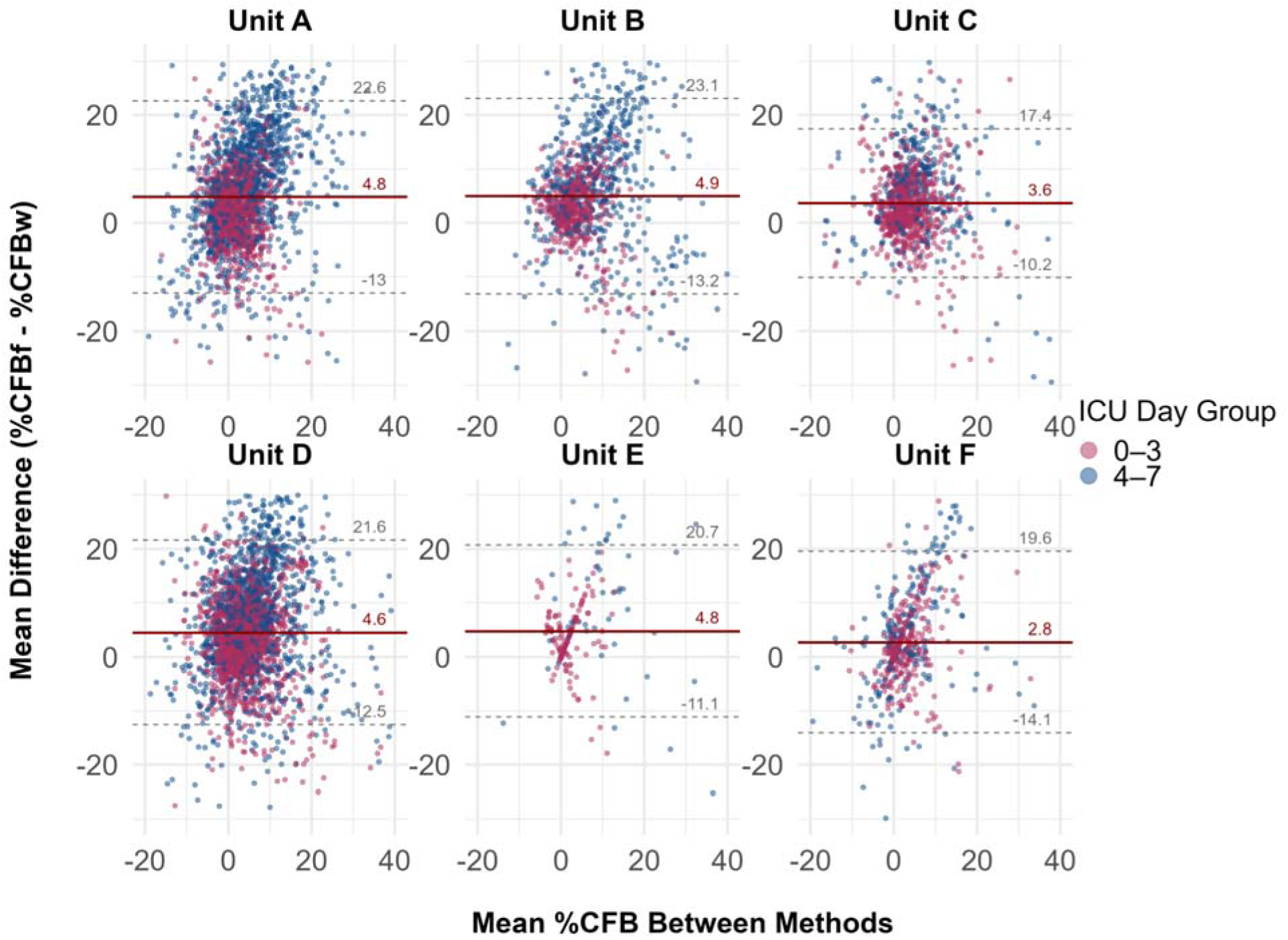
Bland-Altman Plots for all units distinguished by ICU day of CFB calculation. Each point represents the relationship between the two methods for an individual patient. The red horizontal line shows the mean difference between CFBf and CFBw for each unit. The grey dashed lines show the 95% confidence interval for each mean difference. Values with a mean difference greater >30% or <-30%, or with a mean >40% or <-20% were excluded from this figure for visual clarity but were included in the complete analysis.

## RESULTS

We included 9,832 PICU and 2,534 PCICU encounters for 8,895 unique patients, with demographic information listed in **Table 1**. Median (interquartile range, IQR) age of patients at ICU admission varied across units from 0.2 (0 - 2.6) to 7.1 (1.8 - 7.9) years. The proportion of patients who were neonates in each unit varied from 16 - 29% in PCICUs and 1.6 - 2.0% in PICUs. Across all units, 42 - 45% of patients were female, and 42 - 59% of patients were Caucasian. The median (IQR) length of PICU stay was 2 (2–3) days, and the median (IQR) length of PCICU stay was 4.5 (4–5) days.

The ratio of additional weights per ICU encounter, as an estimate of the frequency with which additional weights are measured per ICU encounter, ranged from 0.2 (infrequent) to 1.8 (more frequent), significantly differing across units (**Table 1**, p<0.005). At each unit, we failed to find a significant association between patients’ ventilator status and the amount of additional weights (p>0.10). The proportion of encounters with anchor weights recorded on ICU day 0 ranged from 12% to 62%, significantly differing across units (Table 1, p<0.05). The proportion of encounters with encounter weights obtained after day 0 ranged from 2% to 10%, significantly differing across units (Table 1, p<0.01). Unit C had both the greatest repeated weight frequency (1.8 additional weights per ICU admission) and Unit E had the lowest proportion of ICU patients with anchor weights recorded after ICU day 0 (2%, **Table 1**). At Units B and D, neonates had significantly more additional weights compared to all other age categories (**Supplemental eTable 1,** p<0.001). Additionally, the frequency of encounters with unmeasured urine occurrences ranged from 17% to 78%, significantly differing by unit (p<0.001; **Table 1**). The mean (standard deviation, SD) number of non-anchor weights per patient was higher in the PCICUs (Units A and B: 1.5 (SD 0.4) weights per patient) compared to the PICU units (0.8 (SD 0.6) weights per patient). Additionally, across all sites, there were 1,064 patients (8.6%) with duplicated weights throughout ICU admission (i.e., that varied by less than 0.001 kg to the patient’s anchor weight).

We calculated a total of 8,634 (unit variation: 224 to 3,154) CFB values by each method to match and compare (**Supplemental eTable 1**), determined by the number of additional (new) weights provided by each unit. Both CFBf and CFBw were generally positive throughout ICU admission, with CFBf increasing faster than CFBw (**Figure 1**). Across all units, the mean difference and 95% confidence interval (CI) between CFBf and CFBw were positive (mean differences ranging from 3.6 to 5.3% (CI [3.2, 4.0] to [4.0, 6.5] CFB across units, **Figure 3**), indicating that CFBf was consistently of a greater magnitude than CFBw. When comparing patients with early anchor weights to all patients, Units B, C, and D showed increased mean difference between CFBf and CFBw, but this increased mean difference was only statistically significant at Unit C (**Figure 3 and Supplemental eTable 1**). Similarly, Units A, C, D, and E demonstrated increased mean difference for neonates compared to all patients, though this increase was only statistically significant in Units A and C (**Figure 3**) and was limited by the low number of encounters for neonates in Units E and F.

**Figure 3.**
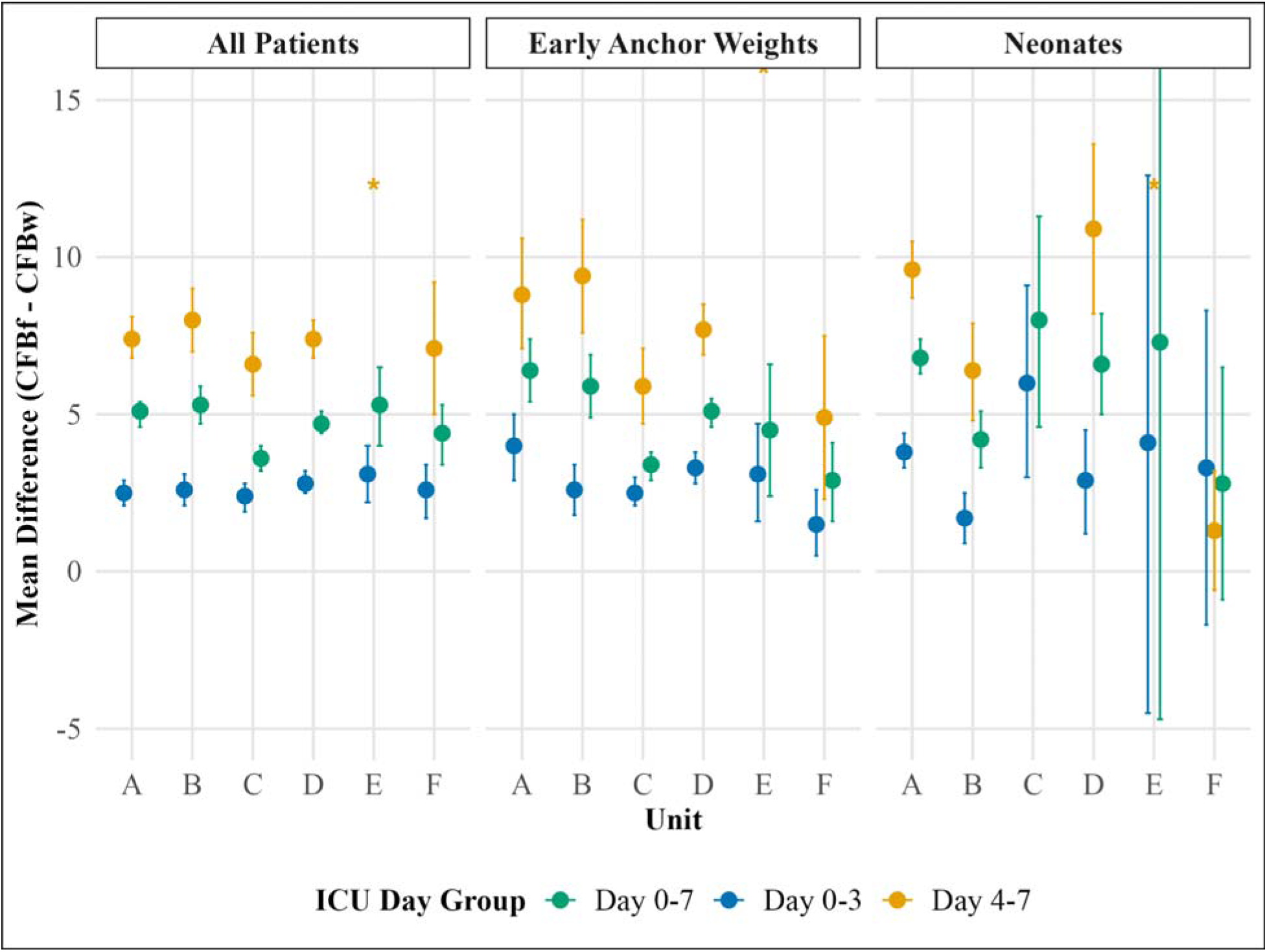
Mean difference between CFBf and CFBw and 95% confidence interval (CI) for each patient subgroup by unit. Calculations shown for ICU days 0-7, further grouped by ICU days 0-3 and ICU days 4-7. Early anchor weight is defined as the first weight recorded on ICU day 0. Asterisks are shown for outliers to improve visual clarity, but were included in the analysis. Unit E, for neonates (days 0-7), exceeds the y-axis limits, with a 95% CI from [-47, 19.3].

### Early versus Late ICU days

When grouping CFB values calculated from ICU days 0-3 and ICU days 4 - 7, the mean difference increased, or CFBf became more divergent from CFBw, across all units (p<0.05, **Figure 3**). A sensitivity analysis focused on encounters with early recorded anchor weights maintained this statistically significant increase in mean difference from ICU days 0 - 3 to ICU days 4 - 7 (p<0.001) at all but Unit F. Neonates also showed a significant difference in the mean difference between CFB methods calculated on ICU days 0 - 3 and ICU days 4 - 7 (p<0.05) across all units.

### Impact of Unmeasured Urine Occurrences

Excluding values calculated after a patient’s first occurrence resulted in a slight decrease, but still positive, mean difference between CFBf and CFBw (**Table 2**). This exclusion reduced available CFB comparisons by between 19% and 68%. All units, except Unit F, showed a decrease in the average mean difference across ICU days 0 – 7 when excluding CFB values calculated after a patient’s first occurrence (mean [SD] decrease for all patients: −0.65 [0.78] vs early anchor weight: −1.2 [1.39] vs neonates: −0.70 [1.01]; **Supplemental eTable 2**). However, this decrease in the difference between CFBf (Pre-Occurrence) and CFBw was only significant for patients with early anchor weights on ICU days 0 - 3 (**Table 2**). Only Unit B showed a significant difference between values calculated up until a patient’s first unmeasured occurrence compared to all values calculated over 7 days of ICU admission (**Table 2**). Both neonates and patients with early anchor weights showed greater mean differences between CFBf and CFBw than CFBf (Pre-Occurrence) and CFBw on ICU days 4 - 7 compared to ICU days 0 - 3 (**Table 2**). All units still had a positive mean difference between CFBf (Pre-Occurrence) and CFBw (**Table 2**).

**Table 2.** Mean difference between CFBf (Pre-Occurrence) and CFBw for different groups with 95% confidence interval. For each row of days (e.g., “Days 0-7”), N gives the number of CFB (Pre-Occurrence) values matched in the Bland-Altman analysis, and (%) represents the proportion of values retained in the Bland-Altman analysis after excluding CFB values calculated after a patient’s first occurrence.

| Unit | A | B | C | D | E | F |
| --- | --- | --- | --- | --- | --- | --- |
| **All Patients (Pre-Occurrence)** |  |  |  |  |  |  |
| **Days 0-7, n (%)** | 1869 (81%) | 364 (33%) | 974 (78%) | 1922 (61%) | 115 (51%) | 323 (56%) |
| Mean Difference | 4.8 [4.3, 5.2] | 2.8 [2.0, 3.6] | 3.2 [2.7, 3.7] | 4.7 [4.2, 5.1] | 4.3 [2.7, 6.0] | 4.6 [3.4, 5.9] |
| **Days 0-3, n (%)** | 1011 (88%) | 262 (47%) | 714 (80%) | 1229 (67%) | 94 (55%) | 224 (64%) |
| Mean Difference | 2.5 [2.0, 2.9] | 1.5 [0.8, 2.3] | 2.1 [1.6, 2.6] | 2.7 [2.3, 3.2] | 2.4 [1.2, 3.5] | 2.7 [1.7, 3.7] |
| **Days 4-7, n (%)** | 858 (74%) | 100 (18%) | 260 (72%) | 693 (53%) | 21 (39%) | 99 (44%) |
| Mean Difference | 7.5 [6.7, 8.2] | 6.0 [3.9, 8.1] | 6.2 [5.0, 7.3] | 8.2 [7.3, 9.0] | 13.3 [7.1, 19.4] | 9.1 [6.0, 12.1] |
| **Early Anchor Weights (Pre-Occurrence)** |  |  |  |  |  |  |
| **Days 0-7, n (%)** | 378 (87%) | 129 (33%) | 540 (78%) | 985 (61%) | 23 (49%) | 91 (56%) |
| Mean Difference | 6.0 [4.9, 7.1] | 2.5 [1.0, 3.9] | 3.1 [2.5, 3.6] | 4.9 [4.3, 5.4] | 2.3 [-0.03, 4.5] | 2.2 [0.7, 3.7] |
| **Days 0-3, n (%)** | 204 (94%) | 97 (32%) | 411 (78%) | 639 (67%) | 23 (55%) | 67 (69%) |
| Mean Difference | 3.8 [2.8, 4.9] | 1.6 [0.4, 2.9] | 2.2 [1.7, 2.7] | 3.1 [2.5, 3.7] | 2.3 [-0.03, 4.5] | 0.9 [-0.3, 2.1] |
| **Days 4-7, n (%)** | 174 (80%) | 32 (17%) | 129 (76%) | 346 (53%) | 0 (0%) | 24 (37%) |
| Mean Difference | 8.6 [6.6, 10.5] | 4.9 [0.8, 9.0] | 5.9 [4.5, 7.3] | 8.2 [7.1, 9.2] | - | 5.8 [1.6, 10.0] |
| **Neonates (Pre-Occurrence)** |  |  |  |  |  |  |
| **Days 0-7, n (%)** | 1078 (86%) | 189 (36%) | 29 (94%) | 173 (79%) | 5 (100%) | 10 (83%) |
| Mean Difference | 6.4 [5.8, 7.0] | 1.7 [0.4, 2.9] | 7.1 [4.0, 10.1] | 7.7 [6.0, 9.3] | 7.1 [-5.1, 19.3] | 1.8 [-2.4, 6.0] |
| **Days 0-3, n (%)** | 568 (94%) | 128 (52%) | 25 (100%) | 95 (81%) | 3 (100%) | 7 (78%) |
| Mean Difference | 3.7 [3.2, 4.3] | 0.7 [-0.6, 1.9] | 6.0 [2.9, 9.0] | 3.7 [2.1, 5.4] | 4.1 [-4.4, 12.6] | 1.3 [-0.6, 3.2] |
| **Days 4-7, n (%)** | 510 (79%) | 6 (22%) | 4 (67%) | 78 (76%) | 2 (100%) | 3 (100%) |
| Mean Difference | 9.4 [8.4, 10.4] | 3.8 [0.9, 6.7] | 13.9 [4.2, 23.7] | 12.4 [9.7, 15.1] | 11.7 [-21.9, 45.2] | 1.3 [-0.6, 3.2] |

## DISCUSSION

Across all patients and units, CFBf was greater in magnitude than CFBw, and differences increased later in ICU admission (days 4 - 7), especially for neonates and patients with early anchor weights (recorded on ICU day 0). Excluding values calculated after a patient’s first unmeasured urine occurrence still maintained a statistically significant mean difference between methods, meaning this difference is likely not solely a result of unmeasured occurrences. To our knowledge, this is the first multicenter, pragmatic observational study to measure these mean differences across children admitted to medical/surgical PICUs and PCICUs over 7 ICU days.

Although there was a relationship between CFBf and CFBw, both methods have inherent clinical complications that might confound the calculation of fluid balance (2). Neonates demonstrate nutritional weight gain during ICU admission, which may be incorrectly attributed to fluid balance (17). Our work, however, showed a greater divergence between CFBf and CFBw for neonates, suggesting neonates’ weight did not increase as quickly as net fluid through ICU day 7. There are also well-documented challenges in obtaining accurate weights for critically ill children, including practical difficulties in measurement and standard error inherent to recording weights (18). In this study, units range from a 0.2 - 1.8 ratio of additional weights per cumulative ICU day, likely reflecting a perceived clinical challenge that varies between units. Additionally, across all units, there were 1,064 patients with weights throughout ICU admission identical to 0.001 kg to the patient’s anchor weight, which might reflect documentation shortcuts. Past research also highlights challenges in charting complete fluid records (19). Inherent to CFBf calculations, a large amount of “insensible” fluid is known to be lost through respiration or perspiration, which is unaccounted for in CFBf calculations (20). Because of these apparent barriers to true CFB assessment, and despite a consistent albeit discrepant relationship between CFBw and CFBf, we cannot confidently determine which method best approximates the patient’s true CFB.

A similar mean difference between CFBf and CFBw has been reported previously during the first 48 hours of PICU admission (13, 21) and over 72 hours in non-critically ill neonates and adults (22, 12) in single-center settings. Similarly, the mean difference between methods increases when anchor weights are taken closer to ICU admission, likely reflecting the positive trend in patient weight from hospital admission through ICU admission (23). Both findings suggest fluid accumulation starts early during hospital and ICU admission, which later anchor weights may fail to capture (24). Because CFBf consistently exceeds CFBw, the relative difference is proportionally larger in neonates and patients with early, and likely lower, anchor weights. With this finding, using an earlier anchor weight to calculate CFB results is likely a more accurate reflection of overall ICU fluid status. In previous studies of patients receiving renal replacement therapy, predicted dry ICU admission weights correlated more closely with organ dysfunction than original anchor weights (25), highlighting the importance of an anchor weight that approximates a patient’s true “dry” weight. Although we are unable to know theoretical dry weight for the patients in our study, the fact that as high as 11% of encounters lacked an early anchor weight suggests that more work can be done to improve the accuracy of our CFB calculations.

An obvious potential source of divergence between CFBf and CFBw is unmeasured output values. In our study, excluding CFB values after a patient’s first unmeasured urine occurrence only contributed to a decrease in the mean difference from 0.65 - 1.2% across all patient groups. Previous studies have quantified the minimum at which differences in fluid measurements are considered clinically significant (13), but inconsistent trends in our units’ relationship between CFBf (pre-occurrence) and CFBf with CFBw limited direct quantitative comparison. No consistent mean difference change was seen on ICU days 4–7, and fewer values were excluded, which may reflect differences for patients with conditions associated with longer ICU stays and more precise urine output tracking (**Table 2**). Some studies have considered stool volume when evaluating fluid output, which we did not do in this study, unless they were measured as a urine/stool mixture with an associated volume; inclusion of this volume would have likely improved agreement between the two CFB methods (27). Importantly, we did not evaluate the contribution of urine occurrences to individual patients’ CFB due to limited weight measurements. Units that had infrequent weight measurements might not have shown a change in mean difference due to fewer weights taken after the patient’s first occurrence.

There were several important limitations to this study. The count of daily weights reported in the dataset limited the comparisons between CFBf and CFBw to a small subset of measures per patient that were done at non-standardized times during the ICU course. As a result, we evaluated differences between values calculated on early versus late ICU days and pre-/post-occurrence on a per-patient basis. Additionally, none of our analyses evaluated the clinical risks associated with increased CFB via each method in our patient population. Due to limitations in data extraction, we had to assume an approximately linear distribution of intake and output subtype across an ICU day. This minimizes errors at weight times close to the beginning and end of each day (7 AM) but maximizes potential errors at weights furthest from this day switch (e.g., 7 PM). Federated analysis did not allow for cross-unit regression comparison, and no severity of illness measures were available. Lastly, we did not correlate either methodology to ICU outcomes, which will be done in future studies to further evaluate which method may be most clinically meaningful.

## CONCLUSION

We report a consistent positive difference between CFB calculated using fluid intake-output versus weights, which generally increased throughout ICU admission. Neonates with relatively small ICU weights and patients with early anchor weights had increased mean differences between methods. Using an early anchor weight, recorded close to ICU admission, also affected this relationship, likely providing a more accurate baseline for change in fluid balance throughout admission. Although urine occurrences contributed to the divergence between these methods, CFBf recorded pre-occurrence was still significantly greater than CFBw across units and patient subgroups. These findings underscore the limitations of both methods and highlight the importance of recording accurate admission weights and frequent weight measurements in critically ill children. Future work must assess the prognostic value of each CFB method in relation to patient outcomes for all critically ill patients, controlling for severity of illness, to better evaluate the clinical significance of the difference between methods.

## Data Availability

All data produced in the present study are available upon reasonable request to the authors

## ACKNOWLEDGEMENTS

The authors would like to acknowledge Sheel Chitre and Adrian La Duca (NYU), Amina Khan (CHOP), and Heather Dutton (UAB) for their assistance with clinical data extraction. We would also like to thank Dr. Benjamin Kozyak, attending physician in the Cardiac Intensive Care Unit at the Children’s Hospital of Philadelphia, for his assistance with CHOP PCICU data analysis.

