## Supplementary Data for "Divergence Between Net Fluid and Weight-Based Evaluation in Calculating Cumulative Fluid Balance"

**TABLE OF CONTENTS:**

- eTable 1: Mean difference between all CFBf and CFBw calculated over PICU admission
- eTable 2: Average difference in mean difference between CFBf and CFBw across units
- eTable 3. Strengthening the Reporting of Observational Studies in Epidemiology (STROBE) Checklist for Cohort Studies.

| **Unit** | **A** | **B** | **C** | **D** | **E** | **F** |
| --- | --- | --- | --- | --- | --- | --- |
| **All Patients** | | | | | | |
| **Days 0- 7, n** | 2308 | 1121 | 1251 | 3154 | 224 | 576 |
| Mean Difference | 5.1 [4.6, 5.4] | 5.3 [4.7, 5.9] | 3.6 [3.2, 4.0] | 4.7 [4.4, 5.1] | 5.3 [4.0, 6.5] | 4.4 [3.4, 5.3] |
| **Days 0- 3, n** | 1145 | 557 | 888 | 1841 | 170 | 349 |
| Mean Difference | 2.5 [2.1, 2.9] | 2.6 [2.1, 3.1] | 2.4 [1.9, 2.8] | 2.8 [2.5, 3.2] | 3.1 [2.2, 4.0] | 2.6 [1.7, 3.4] |
| **Days 4-7, n** | 1163 | 565 | 363 | 1313 | 54 | 227 |
| Mean Difference | 7.4 [6.8, 8.1] | 8.0 [7.0, 9.0] | 6.6 [5.6, 7.6] | 7.4 [6.8, 8.0] | 12.1  [8.4, 15.8] | 7.1 [5.0, 9.2] |
| **Early Anchor Weights** | | | | | | |
| **Days 0-7, n** | 434 | 392 | 692 | 1605 | 47 | 162 |
| Mean Difference | 6.4 [5.4, 7.4] | 5.9 [4.9, 6.9] | 3.4 [2.9, 3.8] | 5.1 [4.6, 5.5] | 4.5 [2.4, 6.6] | 2.9 [1.6, 4.1] |
| **Days 0–3, n** | 216 | 204 | 523 | 953 | 42 | 97 |
| Mean Difference | 4.0 [2.9, 5.0] | 2.6 [1.8, 3.4] | 2.5 [2.1, 3.0] | 3.3 [2.8, 3.8] | 3.1 [1.6, 4.7] | 1.5 [0.5, 2.6] |
| **Days 4–7, n** | 218 | 188 | 169 | 652 | 5 | 65 |
| Mean Difference | 8.8 [7.1, 10.6] | 9.4 [7.6, 11.2] | 5.9 [4.7, 7.1] | 7.7 [6.9, 8.5] | 15.8  [4.9, 26.8] | 4.9 [2.3, 7.5] |
| **Neonates** | | | | | | |
| **Days 0-7, n** | 1250 | 530 | 31 | 219 | 5 | 12 |
| Mean Difference | 6.8 [6.3, 7.4] | 4.2 [3.3, 5.1] | 8.0 [4.6, 11.3] | 6.6 [5.0, 8.2] | 7.3  [-4.7, 19.3] | 2.8 [-0.9, 6.5] |
| **Days 0–3, n** | 602 | 247 | 25 | 117 | 3 | 9 |
| Mean Difference | 3.8 [3.3, 4.4] | 1.7 [0.9, 2.5] | 6.0 [3.0, 9.1] | 2.9 [1.2, 4.5] | 4.1 [-4.5, 12.6] | 3.3 [-1.7, 8.3] |
| **Days 4–7, n** | 648 | 283 | 6 | 102 | 2 | 3 |
| Mean Difference | 9.6 [8.7, 10.5] | 6.4 [4.8, 7.9] | 16.2  [6.2, 26.1] | 10.9  [8.2, 13.6] | 12.1  [-20.5, 44.8] | 1.3 [-0.6, 3.2] |

eTable 1. Mean difference [95% confidence interval] between CFBf and CFBw for different groups and units by Bland-Altman analysis. For each row of days (e.g., “Days 0-7”), N gives the number of CFB values matched. Early Anchor Weights were CFB values calculated with an anchor weight recorded on ICU day 0, and neonates were patients < 1 month. All mean differences were positive, indicating that the calculation of CFB using fluid measurements was of a greater magnitude than the calculation using repeated weight measurements.

|  | **All Encounters** | **Neonates** | **Early Weights** |
| --- | --- | --- | --- |
| **Days 0-7** | -0.65 [-1.47, 0.17] | -0.70 [-2.15, 0.75] | -1.21 [-2.27, -0.15] |
| **Days 0-3** | -0.36 [-0.73, 0.02] | -0.076 [-0.85, 0.69] | -0.54 [-0.82, -0.25] |
| **Days 4-7** | 0.26 [-0.84, 1.36] | -0.42 [-2.76, 1.93] | -0.68 [-2.60, 1.24] |

eTable 2: Average change in mean difference between CFBf and CFBw with 95% confidence interval after excluding values calculated after an encounter’s first urine occurrence. Negative values represent a decrease in the difference between the fluid and weight methods. Averages excluded differences in mean difference from unit E for early weights on days 4-7 and unit A for neonates on ICU days 4-7 due to fewer than 10 comparison points. Similarly, differences in mean difference for units E and F for all neonate differences were excluded.

|  | **Item No** | **Recommendation** | **Page No** |
| --- | --- | --- | --- |
| **Title and abstract** | 1 | (*a*) Indicate the study’s design with a commonly used term in the title or the abstract | 1 |
|  |  | (*b*) Provide in the abstract an informative and balanced summary of what was done and what was found | 2-4 |
| **Introduction** | | | |
| Background/rationale | 2 | Explain the scientific background and rationale for the investigation being reported | 5 |
| Objectives | 3 | State specific objectives, including any prespecified hypotheses | 6 |
| **Methods** | | | |
| Study design | 4 | Present key elements of study design early in the paper | 6 |
| Setting | 5 | Describe the setting, locations, and relevant dates, including periods of recruitment, exposure, follow-up, and data collection | 7 |
| Participants | 6 | (*a*) Give the eligibility criteria, and the sources and methods of selection of participants. Describe methods of follow-up | 7 |
|  |  | (*b*) For matched studies, give matching criteria and number of exposed and unexposed | NA |
| Variables | 7 | Clearly define all outcomes, exposures, predictors, potential confounders, and effect modifiers. Give diagnostic criteria, if applicable | 8 |
| Data sources/ measurement | 8* | For each variable of interest, give sources of data and details of methods of assessment (measurement). Describe comparability of assessment methods if there is more than one group | 7-9 |
| Bias | 9 | Describe any efforts to address potential sources of bias | 7 |
| Study size | 10 | Explain how the study size was arrived at | 7 |
| Quantitative variables | 11 | Explain how quantitative variables were handled in the analyses. If applicable, describe which groupings were chosen and why | 9 |
| Statistical methods | 12 | (*a*) Describe all statistical methods, including those used to control for confounding | 9 |
|  |  | (*b*) Describe any methods used to examine subgroups and interactions | 9 |
|  |  | (*c*) Explain how missing data were addressed | NA |
|  |  | (*d*) If applicable, explain how loss to follow-up was addressed | NA |
|  |  | (*e*) Describe any sensitivity analyses | 9 |
| **Results** | | |  |
| Participants | 13* | (a) Report numbers of individuals at each stage of study—eg numbers potentially eligible, examined for eligibility, confirmed eligible, included in the study, completing follow-up, and analysed | NA |
|  |  | (b) Give reasons for non-participation at each stage | NA |
|  |  | (c) Consider use of a flow diagram | NA |
| Descriptive data | 14* | (a) Give characteristics of study participants (eg demographic, clinical, social) and information on exposures and potential confounders | 9 |
|  |  | (b) Indicate number of participants with missing data for each variable of interest | NA |
|  |  | (c) Summarise follow-up time (eg, average and total amount) | NA |
| Outcome data | 15* | Report numbers of outcome events or summary measures over time | 10-11 |

| Main results | 16 | (*a*) Give unadjusted estimates and, if applicable, confounder-adjusted estimates and their precision (eg, 95% confidence interval). Make clear which confounders were adjusted for and why they were included | 10-12 |
| --- | --- | --- | --- |
|  |  | (*b*) Report category boundaries when continuous variables were categorized | NA |
|  |  | (*c*) If relevant, consider translating estimates of relative risk into absolute risk for a meaningful time period | NA |
| Other analyses | 17 | Report other analyses done—eg analyses of subgroups and interactions, and sensitivity analyses | 11 |
| **Discussion** | | | |
| Key results | 18 | Summarise key results with reference to study objectives | 12 |
| Limitations | 19 | Discuss limitations of the study, taking into account sources of potential bias or imprecision. Discuss both direction and magnitude of any potential bias | 14-15 |
| Interpretation | 20 | Give a cautious overall interpretation of results considering objectives, limitations, multiplicity of analyses, results from similar studies, and other relevant evidence | 12-14 |
| Generalisability | 21 | Discuss the generalisability (external validity) of the study results | 13-14 |
| **Other information** | | | |
| Funding | 22 | Give the source of funding and the role of the funders for the present study and, if applicable, for the original study on which the present article is based | 1 |

.

eTable 3. Strengthening the Reporting of Observational Studies in Epidemiology (STROBE) Checklist for Cohort Studies. Give information separately for exposed and unexposed groups. Numbered by page number in manuscript.
